# Genetic Susceptibility Of Cytotoxic T Lymphocyte-Associated Antigen 4 Gene Polymorphism In The Onset Of Arthritis

**DOI:** 10.1101/2021.04.27.21255970

**Authors:** Maryam Mukhtar, Nadeem Sheikh, Saira Kainat Suqaina, Tayyaba Saleem, Rabia Mehmood, Muhammad Babar Khawar

**Author notes:** **Corresponding Author** E-mail address, Mailing address: Cell and Molecular Biology Lab, Department of Zoology, University of the Punjab, Quaid-i-Azam Campus, Lahore, 54590.

## Abstract

Cytotoxic T lymphocyte-associated antigen 4 (CTLA-4) gene plays a vital role in the activation of T-cells as a down regulator. CTLA-4 gene polymorphisms have implicated a potential risk factor for autoimmune disorders like arthritis. Therefore the current study was designed to determine the association of CTLA-4 gene polymorphism in the onset of rheumatoid and osteoarthritis in Pakistani individuals. Genotyping was performed on 300 RA, 316 OA, and 412 control subjects by direct sequencing method as well as polymerase chain reaction-restriction fragment length polymorphism (PCR-RFLP) technique. It was observed that allelic and genotypic frequency of rs5742909, rs231775, rs4553808, rs733618, and rs3087243 were significantly varied among patients and controls and considered as a significant risk factor in the onset of RA as well as OA. However, no mutation was identified on the rs11571317 polymorphic site. Haplotype CAGTCA and CAG TCG act as a protectant against disease onset whereas CAACCG was significant in disease onset. Mutation on rs231775 polymorphic site lead to the change of threonine into alanine It was concluded that CTLA-4 gene polymorphism is a significant risk factor in the onset of RA as well as OA. Large scale survey is required for the screening of the genetic markers for pre-diagnosis of the disease.

**SUMMARY STATEMENT:** The study summarized that CTLA-4 gene polymorphism plays a key role in the arthritis onset in Pakistani population.

## Introduction

An inflammatory disorder named arthritis affects multiple body joints with varying factors like infections, trauma, autoimmune disorders, aging and idiopathic causes (1). It has multiple types whereas two were of major i-e., Rheumatoid arthritis (RA) and Osteoarthritis (OA). A systematic disease RA is characterized by debilitating, destructive and chronic arthritis (2). However OA is changes in articular cartilage, related changes in joint margins, underlying bones (3).

Arthritis is polygenic disorder and caused by interaction of mutations on multiple genes like *PADI-4, PTPN22* and *CTLA-4* (4;5). Cytolytic T lymphocyte-associated antigen 4 (*CTLA-4*) negatively regulate responses of immuno T cells and vital checkpoint in altering antitumor responses and autoimmunity. Polymorphisms on *CTLA-*4 genes in humans altered *CTLA-4* that ultimately lead to the immune dysregulation syndromes onset (6).

*CTLA-4* gene was located on chromosome 2q33 comprised of a leader sequence and 3 exons as well as exist as monocopy per haploid genome (7;8). Three polymorphism sites were reported on *CTLA-4* locus. Out of which first polymorphism was the dinucleotide i-e., (AT)n repeat at 642bp of exon 3. The second polymorphic site was transition of G to A at 49 (G49A) position of exon 1(9). While the third was first reported by (7), and of C to T transition of promotor sequence at −318 (C-318T) position.

Pakistan because of intracaste marriages is at higher risk of genetic disorders including arthritis which is the leading cause of disability. With increasing age the risk of disease development also increasing. However the basic genetic factors associated with arthritis is still unknown. There for the aim and objective of present study is to determined the susceptible association of *CTLA-4* gene aith the onset of RA as well OA on studied population.

## Result

As a result of genotyping it was observed that polymorphism exist on all targeted polymorphic sites on *CTLA-4* gene except rs11571317. On rs5742909 site, T allele was more prevalent among patients as compared to C allele. Similarly G allele was more prevalent on rs231775 and rs4553808 among patients instead of allele A. C allele was replaced by T allele on rs733618 and on rs3087243 polymorphic site allele G was more common among patients as compared to controls. All SNP’s followed HWE (p > 1.00). The allelic and genetic analysis was performed and was presented in table 2 and 3. No mutation was observed on rs11571317 polymorphic site. The allelic frequency of mutant allele on rs231775, rs733618 and rs3087243 were significantly varied among RA patients and control. However in OA patient’s rs5742909, rs231775, rs4553808 and rs3087243 were significantly varied in comparison to controls (p < 0.01). At genetic level, it was observed that all SNP’s except rs5742909 were significantly associated with the onset of RA (p < 0.01) where as in OA rs733618 was not significantly associated with onset of disease (p<0.01).

**Table 1:**
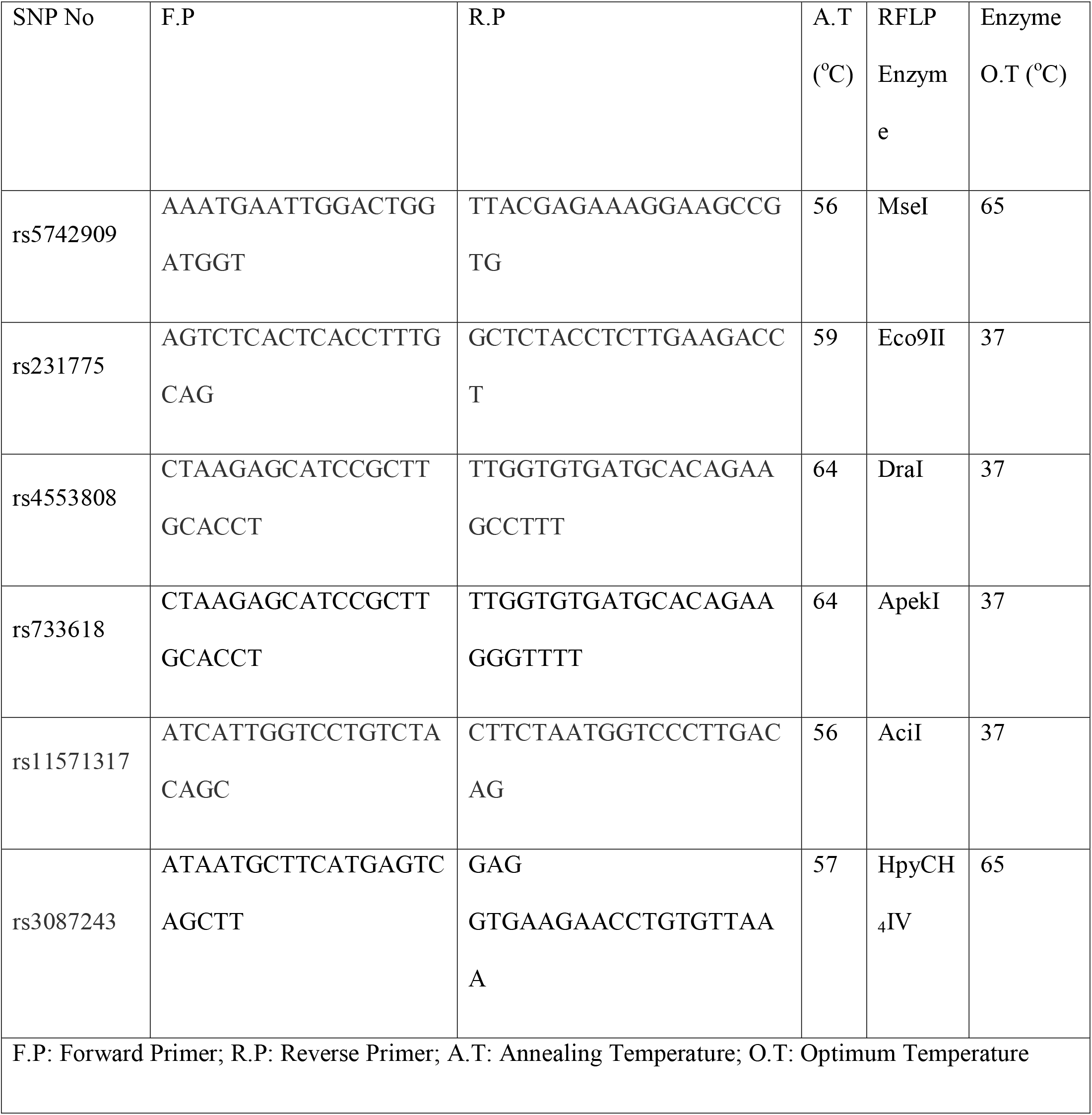
PCR and RFLP enzymes conditions for selected SNP’s on *CTLA-4* gene.

**Table 2.** Allelic test of genetic variants of *CTLA-4* gene in RA, OA and controls.

| SNP's | Alleles | RA | p-value | OA | p-value |
| --- | --- | --- | --- | --- | --- |
|  |  | Frequency<br>(P/C) |  | Frequency<br>(P/C) |  |
| rs5742909 | C | 0.302/1.000 | <0.01 | 0.419/0.164 | 0.016* |
|  | T | 0.698/0.000 |  | 0.581/0.836 |  |
| rs231775 | A | 0.225/1.000 | 0.017* | 0.217/1.000 | 0.015* |
|  | G | 0.775/0.000 |  | 0.581/0.000 |  |
| rs4553808 | A | 0.737/0.000 | <0.01 | 0.828/0.038 | 0.016* |
|  | G | 0.263/1.000 |  | 0.172/0.962 |  |
| rs733618 | C | 0.920/0.000 | 0.015* | 0.540/0.000 | <0.01 |
|  | T | 0.080/1.000 |  | 0.460/1.000 |  |
| rs11571317 | C | 1.000/1.000 | - | 1.000/1.000 | - |
| rs3087243 | A | 0.457/0.917 | 0.015* | 0.055/0.917 | 0.016* |
|  | G | 0.543/0.083 |  | 0.945/0.083 |  |

\* Represents significance at the 0.01 level.

**Table 3.** Genetic test of genetic variants of *CTLA-4* gene in RA, OA and controls.

| **SNP's** | **Genotypes** | **RA**<br>**Frequency**<br>**(P/C)** | **p-**<br>**value** | **OA**<br>**Frequency**<br>**(P/C)** | **p-value** |
| --- | --- | --- | --- | --- | --- |
| rs5742909 | CC | 0.067/1.000 | 0.016* | 0.089/1.000 | <0.01 |
|  | CT | 0.470/0.036 |  | 0.661/0.000 |  |
|  | TT | 0.463/0.964 |  | 0.250/0.00 |  |
| rs231775 | AA | 0.040/1.000 | 0.015* | 0.038/1.000 | 0.132 |
|  | AG | 0.370/0.000 |  | 0.358/0.000 |  |
|  | GG | 0.590/0.000 |  | 0.604/0.002 |  |
| rs4553808 | CC | 0.473/0.000 | 0.016* | 0.828/0.000 | 0.001* |
|  | CT | 0.527/0.000 |  | 0.010/0.000 |  |
|  | TT | 0.000/1.000 |  | 0.162/1.000 |  |
| rs733618 | CC | 0.840/0.000 | <0.01 | 0.079/0.000 | 0.015* |
|  | CT | 0.160/0.000 |  | 0.921/0.000 |  |
|  | TT | 0.000/1.000 |  | 0.000/1.000 |  |
| rs11571317 | CC | 1.000/1.000 | - | 1.000/1.000 |  |
| rs3087243 | AA | 0.097/0.835 | 0.015* | 0.089/0.935 | 0.001* |
|  | AG | 0.720//0.165 |  | 0.730/0.065 |  |
|  | GG | 0.183/0.000 |  | 0.183/0.000 |  |

\* Represents significance at the 0.05 level.

Linkage disequilibrium analysis was presented in Figure 1 (a, b) in RA patients and Figure 2 (a, b) in OA patients. In comparison of controls, in RA rs5742909 with rs231775 were higher risk factor (D’=0.975; r2=0.787) followed by rs11571317 along with rs5742909 and rs231775 (D’=0.840, r2=0.465) and (D’=0.831, r2=0.529) respectively. Whereas variants on rs231775, rs4553808 and 733618 in combination were increased chances on RA development. On other hand, in OA subjects and controls if all SNP’s will forward to next generation they all in combination increased chances of disease onset (D’=1.000, r2=0.401). if rs733618, rs11571317 and rs3087243 were significant risk factor in combination (D’=0.914, r2=0.512).

**Figure 1.**
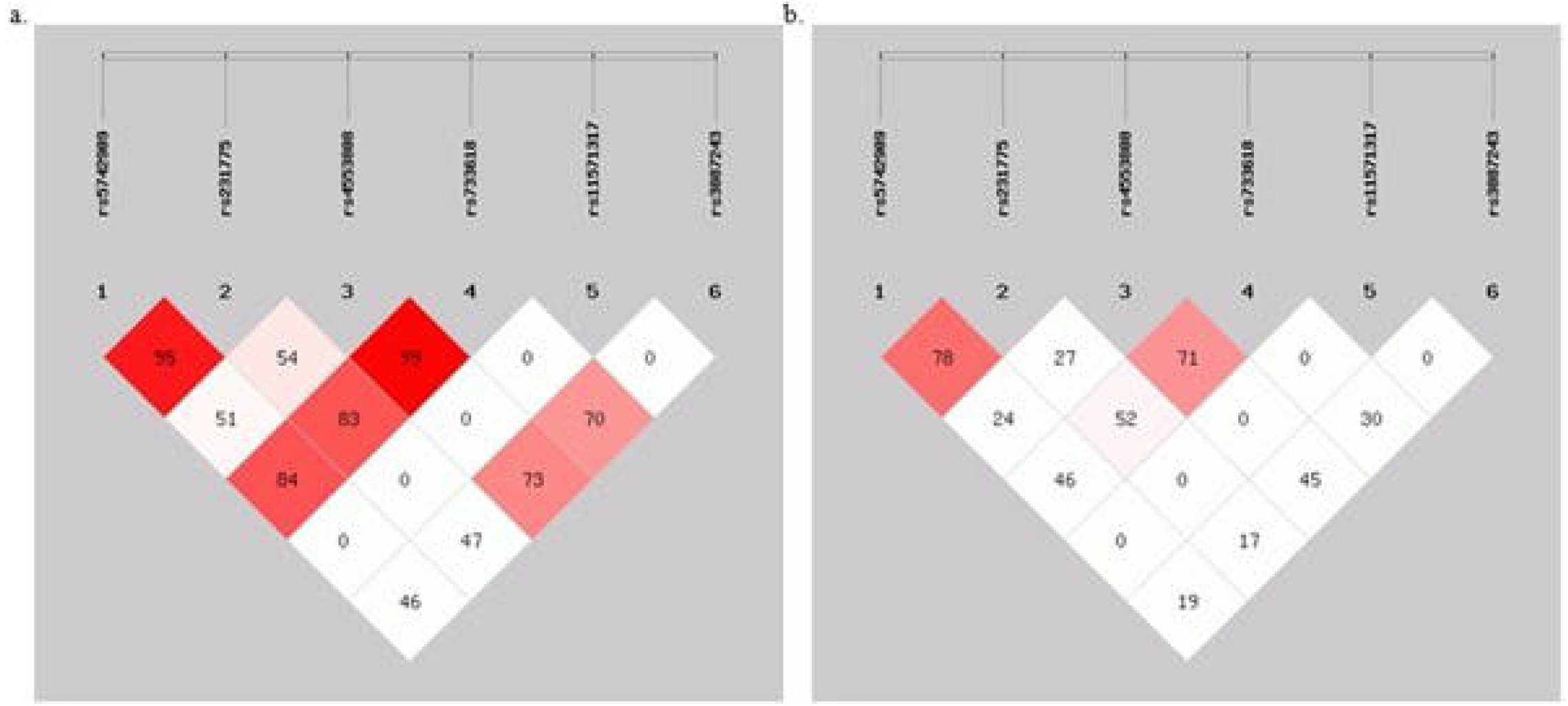
Location and map of Linkage Disequilibrium (LD) in SNPs at *CTLA-4* gene in RA are presented. The SNPs numbers are indicated at the top of haploview. (a) LD = D^/^ (b) r^2^ = LD Coefficient.

**Figure 2.**
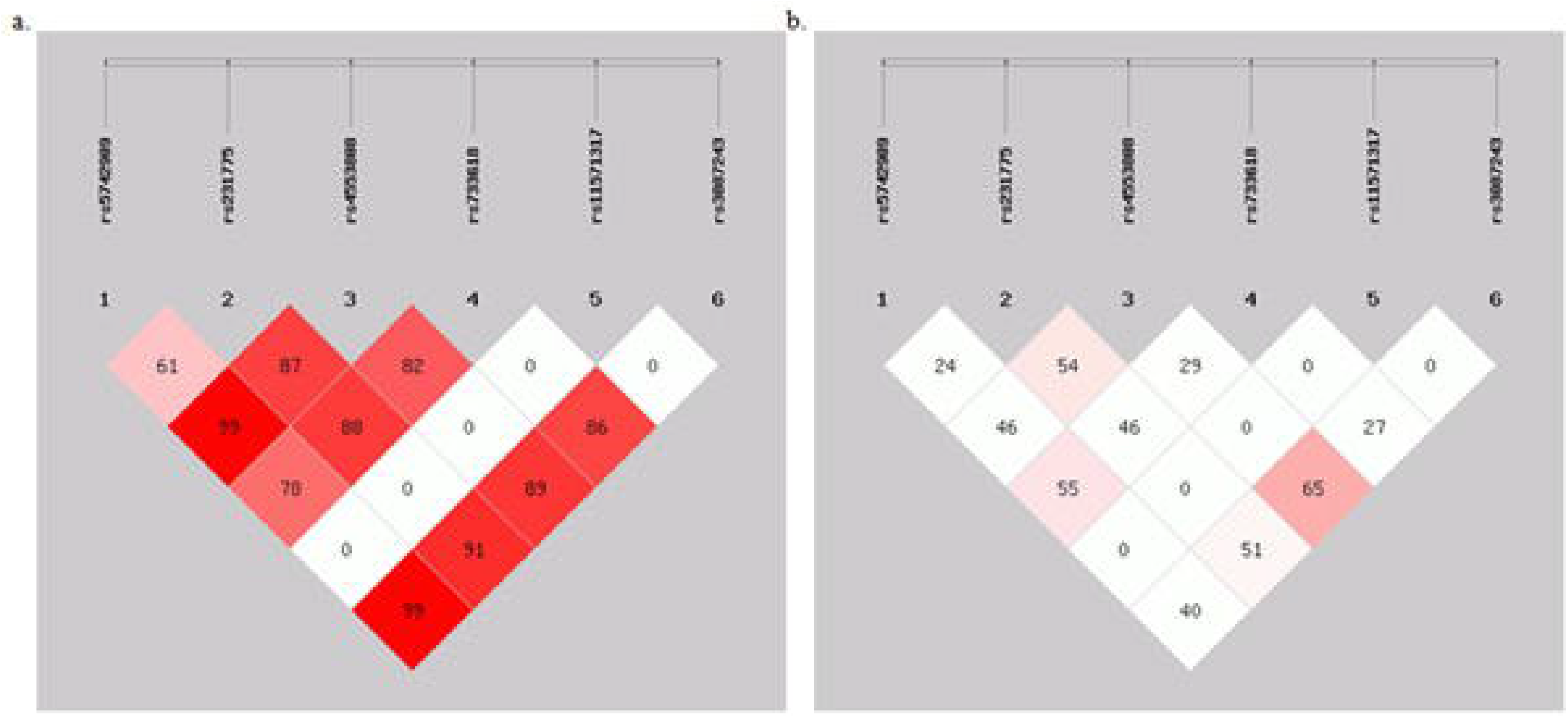
Location and map of Linkage Disequilibrium (LD) in SNPs at *CTLA-4* gene in OA are presented. The SNPs numbers are indicated at the top of haploview. (a) LD = D^/^ (b) r^2^ = LD Coefficient.

It was observed that in RA and OA subjects, the frequency of haplotypes CAGTCA and CAGTCG was higher in controls as compared to patients therefore they were protective against disease onset. The haplotypes significant in disease onset were summarized in Table 4.

**Table 4.** Haplotype Analysis of the *CTLA-4* gene located on chromosome 2.

| **Haplotype:** rs5742909, rs231775, rs4553808, rs733618, rs11571317, rs3087243 |  |  |  |  |  |
| --- | --- | --- | --- | --- | --- |
| **Haplotype** | **RA**<br>**Frequency**<br>**(P/C)** | **p-value** | **Haplotype** | **OA**<br>**Frequency**<br>**(P/C)** | **p-value** |
| CAGTCA* | 0.000/0.917 | 0.003 | CAGTCA<br>* | 0.000/0.917 | 0.002 |
| CAGTCG* | 0.000/0.083 | <0.01 | CAGTCG<br>* | 0.000/0.083 | 0.001 |
| CAACCA | 0.104/0.000 | 0.021 | CAACCA<br>* | 0.002/0.000 | 0.005 |
| CAACCG* | 0.099/0.000 | 0.005 | CGACCA<br>* | 0.004/0.000 | 0.003 |
| CGACCA* | 0.026/0.000 | 0.006 | CGACCG<br>* | 0.034/0.000 | 0.005 |
| CAACCG* | 0.041/0.000 | 0.009 | CGGCCG<br>* | 0.123/0.000 | 0.003 |
| CGGCCA* | 0.020/0.000 | 0.005 | CGGTCG<br>* | 0.002/0.000 | 0.002 |
| CGGTCA* | 0.012/0.000 | 0.001 | TAGCCG | 0.198/0.000 | 0.002 |
|  |  |  | * |  |  |
| TAACCG* | 0.023/0.000 | 0.005 | TGGCCG<br>* | 0.024/0.000 | 0.006 |
| TGACCA* | 0.063/0.000 | 0.001 | TGGTCG | 0.359/0.000 | 0.021 |
| TGACCG* | 0.381/0.000 | 0.002 |  |  |  |
| TGGCCA* | 0.163/0.000 | 0.002 |  |  |  |
| TGGTCA* | 0.068/0.000 | 0.001 |  |  |  |

\*Represents a significant association of Haplotypes with Arthritis onset.
⦿ Represents a significant association of Haplotypes protective against Arthritis onset

As a result of protein alignment on MEGA6 software, it was observed that polymorphism on rs231775 lead to the change of threonine into alanine.

## Discussion

During various phases of T cells response, *CTLA-4* performed different independent effects such as apoptosis induction in activated T cells as well as to suppress proliferation of T cells or *CTLA-4* may directly regulate B cells responses (10). The current study was designed with an aim to investigate the susceptibility of C*TLA-4* gene functional polymorphisms with the onset of arthritis.

Multiple SNP’s were identified on studied gene out of which some variants were reported to be associated with the onset of autoimmune disorders like rheumatoid arthritis and Sjogren’s Syndrome (11-13). Current study demonstrated that except rs11571302, all other studied SNP’s including rs5742909, rs231775, rs4553808, rs733618 and rs3087243 polymorphic sites were significantly associated with the onset of RA as well as OA. Similarly rs5742909 and rs231775 were reported as significant susceptible genetic markers in onset of RA (14-16). Similarly rs3087243 was reported as significant risk factor in the onset of RA in Mexican, Chinese and North American populations (17-19). Contrary to this polymorphism on rs5742909 was reported as non significant factor in the onset of RA in Mexican, Spanish and Korean populations (19-21). Rs231775 was also reported as non-significant factor in the onset of RA in british population (5;22;23).

Currently studied polymorphisms were already reported as functional and involved in altering immune response. Mutant allele T on rs5742909 polymorphic site was reported to be involved in elevating activity of promotor which ultimately increased expression of gene and protein of *CTLA-4* mRNA and membrane *CTLA-4* respectively. It also suppressed immune response by negatively regulate T cells (24;25).

Whereas polymorphism on rs231775 site was involved in altering activation of T cells and *CTLA-4* inhibitory function. Allele G on above mentioned polymorphic site lowered protein expression which decreased control on T cells activation and proliferation lead to the onset of autoimmune disorders including RA by altering T cells regulation (14;24;26-28). It was postulated that mutation on rs231775 may also influenced surface trafficking and endocytosis, glycosylation of *CTLA-4* and intracellular portioning which ultimately affected its inhibitory action (29).

Polymorphism on promotor region (rs733618 and rs4553808) of reported to be involved in alternative abnormal slicing which affects gene expression and hence contribute in RA and OA pathogenesis (30)

In pipeline of this, the mutant G allele on rs3087243 polymorphic site altered level of cytoplasmic and membrane CTLA-4 along with sCTLA-4 mRNA and sCTLA-4 levels (31;32).

Current study demonstrated haplotype CAGTCA and CAGTCG were protective against disease onset in Pakistani population where as rest haplotypes were significant risk factors in the onset of disease. Up to best of my knowledge current study is the first ever study reporting the association of functional polymorphic sites of *CTLA-4* gene with RA as well as OA in Pakistani population.

In conclusion it was found that *CTLA-4* is a significant risk factor in the onset of not only RA as well OA. However a large scale screening must be needed so that the above mentioned genetic markers can be used for the pre diagnosis of the disease.

## Materials and Methods

### Sampling

For meeting the above mentioned objective a case control study was designed and ethically approved by the Punjab University Advanced Studies and Research Board, Lahore, Pakistan. The study was recruited from the rheumatology and orthopedic center of Public and Semi Government Hospitals of Punjab, Pakistan. Already diagnosed individuals with disease were selected after taking written consent. The study was comprised of 300 RA, 316 OA and 412 controls subjects. All RA subjects were with positive Rf factor where as OA were with negative Rf Factor. All control subjects’ age and sex matched to the cases and were healthy as well as with negative family history of arthritis.

### Genotyping

DNA was isolated by manually from each blood sample and was stored at -80°C. Genotyping was performed for selected polymorphic sites by sanger’s sequencing method (S. Figure 1) as well as PCR-RFLP technique (S. Figure 2). The primer with respect to the targeted polymorphic sites along with their respective endonucleases enzymes (Thermo Scientific) and conditions was summarized in table 1.

### Statistical Analysis

For genetic analysis whole data was passed through Hardy-Weinberg Equilibrium (p> 0.05). The Fisher’s P test was used for allelic and genotypic frequencies. Linkage disequilibrium level and haplotype were calculated to study the studied SNP’s association with arthritis by SHEsis (http://analysis.bio-x.cn/SHEsisMain.htm). Alterations in amino acid sequences were determined by Mega 6 software.

## Data Availability

The data will be available by email directly to the author.

## Acknowledgement

Authors are thankful to Higher Education Commission and worthy Vice Chancellor, University of the Punjab, and Lahore for providing funding for accomplishment of research.

